# Cerebrospinal Fluid and Plasma Procalcitonin for the Diagnosis of Neonatal Bacterial Meningitis

**DOI:** 10.1101/2021.05.13.21257154

**Authors:** Sourabh Dutta, Naresh Sachdeva, Vilay Sanger, Arnab Pal, Pallab Ray

## Abstract

**Objective:** To determine accuracy of cerebro-spinal fluid (CSF) procalcitonin (PCT) to diagnose neonatal bacterial meningitis (NBM) among septic neonates and compare with other index tests.

**Design:** Prospective, cross-sectional, single-gate study

**Setting:** Level-3 neonatal unit

**Patients:** Neonates with suspected sepsis undergoing lumbar puncture

**Index tests:** CSF PCT, leukocyte count and biochemistry; plasma PCT and CSF:plasma PCT ratio

**Reference standards:** “Definite meningitis” defined by positive CSF culture and/or gram stain and/or broad-based primer 16S rDNA polymerase chain reaction. “Definite or probable meningitis” defined by definite meningitis or probable meningitis (based on cytochemistry cut-offs).

**Results:** Of 216 eligible neonates, 18 had “definite meningitis” and 37 “definite or probable meningitis”. Median (Q_1_, Q_3_) CSF PCT level was higher in “definite meningitis” compared to “no definite meningitis” [0.429 (0.123, 1.300) vs. 0.181 (0.119, 0.286) ng/ml respectively, p=0.028]. Likewise, it was higher in “definite or probable meningitis” compared to no meningitis [0.245 (0.136, 0.675) vs. 0.170 (0.116, 0.28), p=0.01]. The area under ROC curve (AUC) of CSF PCT level for definite meningitis was 0.656 and for “definite or probable meningitis” 0.635. Paired comparisons of AUC of CSF PCT with other index tests were not significant. Based on *a priori* cut-off of 0.2 ng/ml, CSF PCT level had a sensitivity (95% CI) of 67% (50, 80), specificity 58% (54, 61), LR^+^ 1.6 (1.1, 2.0) and LR^-^ 0.6 (0.3, 0.9).

**Conclusions:** Higher values of CSF PCT are associated with NBM. CSF PCT cannot reliably discriminate between meningitis and no meningitis and is not superior to other CSF tests.

## Introduction

Meningitis is more common in neonates than at any other age.^1^ The incidence of neonatal bacterial meningitis (NBM) ranges from 0.21 per 1000 livebirths in developed countries to 6.1 per 1000 livebirths in developing countries.^2^ NBM must be quickly and accurately diagnosed as under-treatment can have long-term neurological consequences.^3^

The clinical signs of NBM are non-specific and overlap with viral meningitis and sepsis. The common reference standard tests are based on demonstrating the etiologic organisms, viz. cerebro-spinal fluid (CSF) culture, gram stain, and polymerase chain reaction (PCR), but none of them is ideal. CSF cultures are often false negative due to prior antibiotic exposure, small sample volumes, and delayed processing.^4^ In clinical practice, most neonates receive antibiotics before a lumbar puncture (LP).^5^ It takes ≥24-48 hours for CSF cultures to be reported positive.^6^ CSF gram stain has limited sensitivity.^7^ In a meta-analysis on PCR-based diagnosis of sepsis in neonates, molecular methods did not have sufficient accuracy to replace microbial cultures.^8^ The sensitivity of broad range PCR to diagnose meningitis is only 86%.^9^ Moreover, broad-based PCR methods are not widely available. Given the limitations with the reference standards, clinicians often rely on CSF white blood cell count (WBCC), protein, and glucose to diagnose meningitis. However, the sensitivity and specificity of these tests, individually and in combination, are unsatisfactory.^10, 11^ Up to 50% of LPs in neonates are traumatic, adding to difficulties in interpretation.^12^ Hence, there is a need for newer and better tests to diagnose NBM.

Procalcitonin (PCT), an inflammatory biomarker, is secreted by the C-cells of the thyroid gland and the neuroendocrine cells of the lung and intestine.^13^ It is an early marker of neonatal sepsis and is widely used to guide antibiotic therapy.^14-16^ There is a physiological postnatal rise in PCT levels which declines and stabilises by about 72-84 hours of life.^17^

In adults, serum PCT distinguishes bacterial from viral meningitis and is superior to serum CRP.^18, 19^ Wei et al. evaluated studies on CSF and serum PCT (including 8 Pediatric studies) to diagnose meningitis. The pooled area under the receiver operator characteristics curve (AUC) for CSF and serum PCT were 0.9 and 0.98 respectively.^20^ The reasons behind high serum PCT level in meningitis and the source of the CSF PCT are not clear from existing literature.

Serum PCT has been extensively studied in neonatal sepsis, but to the best of our knowledge, only two studies have evaluated CSF PCT to diagnose NBM.^21-23^ We hypothesized that among neonates who undergo an LP as part of the sepsis workup, CSF PCT has a greater accuracy and will replace CSF WBCC, biochemistry, plasma PCT for diagnosing definite meningitis.

## Methods

We conducted a prospective, cross-sectional diagnostic accuracy study in a level III neonatal intensive care unit from 2017 to 2020. The study had a single-gated design. We included consecutive neonates of any gestation aged 0-56 days with clinically suspected sepsis as per the NEO-KISS criteria, in whom a lumbar puncture (LP) was planned.^24^ We excluded neonates who received multiple doses of antibiotics before the LP. Post-enrolment, we excluded neonates with an inadequate or macro-traumatic CSF sample, or contaminated CSF culture (aerobic spore bearers, Diphtheroides, or Propionibacterium). We obtained written informed consent from parents of eligible neonates.

We recorded demographic information, risk factors and clinical features. We analyzed the CSF sample for total leukocytes and neutrophil count, glucose, protein, gram stain, and culture by standard methods. 800 µL sample of CSF was frozen at −80°C for real-time PCR. DNA was isolated from CSF using a QIAamp DNA Micro Kit (Qiagen, Germany). We amplified the DNA using broad-range forward and reverse primers: 341F (5’ CCTACGGGNGGCWGCAG 3’) and 785R (5’-GACTACHVGGGTATCTAATCC 3’) in a real-time PCR system (QuantStudio™, Thermo Fisher, USA). The 444 bp-sized amplicon covered the V3-V4 regions of the 16S rDNA gene.

We used 200 µL of CSF for the PCT assay. In addition, we collected an EDTA blood sample within ± 3 hours of performing the LP and used 200 µL plasma for PCT assay. Samples for CSF and plasma PCT were either analysed immediately or frozen at −20°C for upto 24-48 hours before analysis. PCT concentrations were determined by a two-step sandwich electrochemiluminescence assay (Roche Diagnostics, Germany). The assay’s measuring range is 0.02-100 ng/ml with a functional sensitivity of 0.06 ng/ml and analytical sensitivity of 0.02 ng/ml.

Our reference standard was “definite meningitis”, defined as positive CSF culture and/or gram stain and/or PCR. Our primary index test was CSF PCT. We compared CSF PCT against CSF WBCC, neutrophil count, glucose and protein, and plasma PCT and CSF:plasma PCT ratio. In a secondary analysis, the reference standard was “definite or probable meningitis”, with probable meningitis defined as CSF WBCC ≥25 leukocytes with ≥60% neutrophils and/or glucose <20 mg/dL and/or protein >150 mg/dL among term or >180 mg/dL among preterm infants. Since CSF cytochemistry was included in the definition of “definite or probable meningitis”, we restricted the comparison of CSF PCT to plasma PCT and CSF:plasma PCT ratio in the secondary analysis. Personnel performing the index and reference standard tests were blinded to each other and the patient’s clinical status. The results of the index tests did not influence the decision to perform the reference standard tests.

We described and compared data between groups by standard methods. We determined the area under the receiver operator characteristics curve (AUC) for each index test and derived cut-off values based on Youden’s index. We compared the AUCs using a paired non-parametric test. Since plasma PCT has a surge in the first 72 hours of life, we compared CSF PCT in LP’s performed at <72 hours of life versus ≥72 hours. It is unclear how the authors of the previous studies on NBM derived cut-off values for CSF PCT.^22, 23^ Nevertheless, using 0.2 ng/ml as the a priori cut-off value of CSF PCT based on Rajial et al., we determined its sensitivity, specificity, and likelihood ratios (LRs). We compared the correlation coefficients of CSF and plasma PCT levels at <72 hours versus ≥72 hours. Likewise, we compared the correlation coefficients in the “definite or probable meningitis” versus no meningitis groups. We planned to exclude subjects with indeterminate or missing data from the analysis. We did not impute missing data.

Assuming definite meningitis occurred in 8.5% patients with suspected sepsis (unit data), we required 220 subjects to detect sensitivity of 95% with a 10% width of 95% confidence interval (CI). This sample size could detect a specificity of 95% with a 3% width of 95% CI.

The Institute Ethics Committee approved the protocol (PGI/IEC/2015/891 dated 10-2-15).

## Results

We screened 392 neonates, of which 216 were available for analysis (Figure 1). 18 (8.3%) had definite meningitis, of which 17 were CSF culture/gram stain positive, and all were PCR-positive. Thirty-seven neonates had “definite *or* probable meningitis”. 8 met the criteria for definite *and* probable meningitis, and 18 had *only* probable meningitis. *Acinetobacter baumannii* (n=4) was the commonest organism grown from CSF samples.

**Figure 1.**
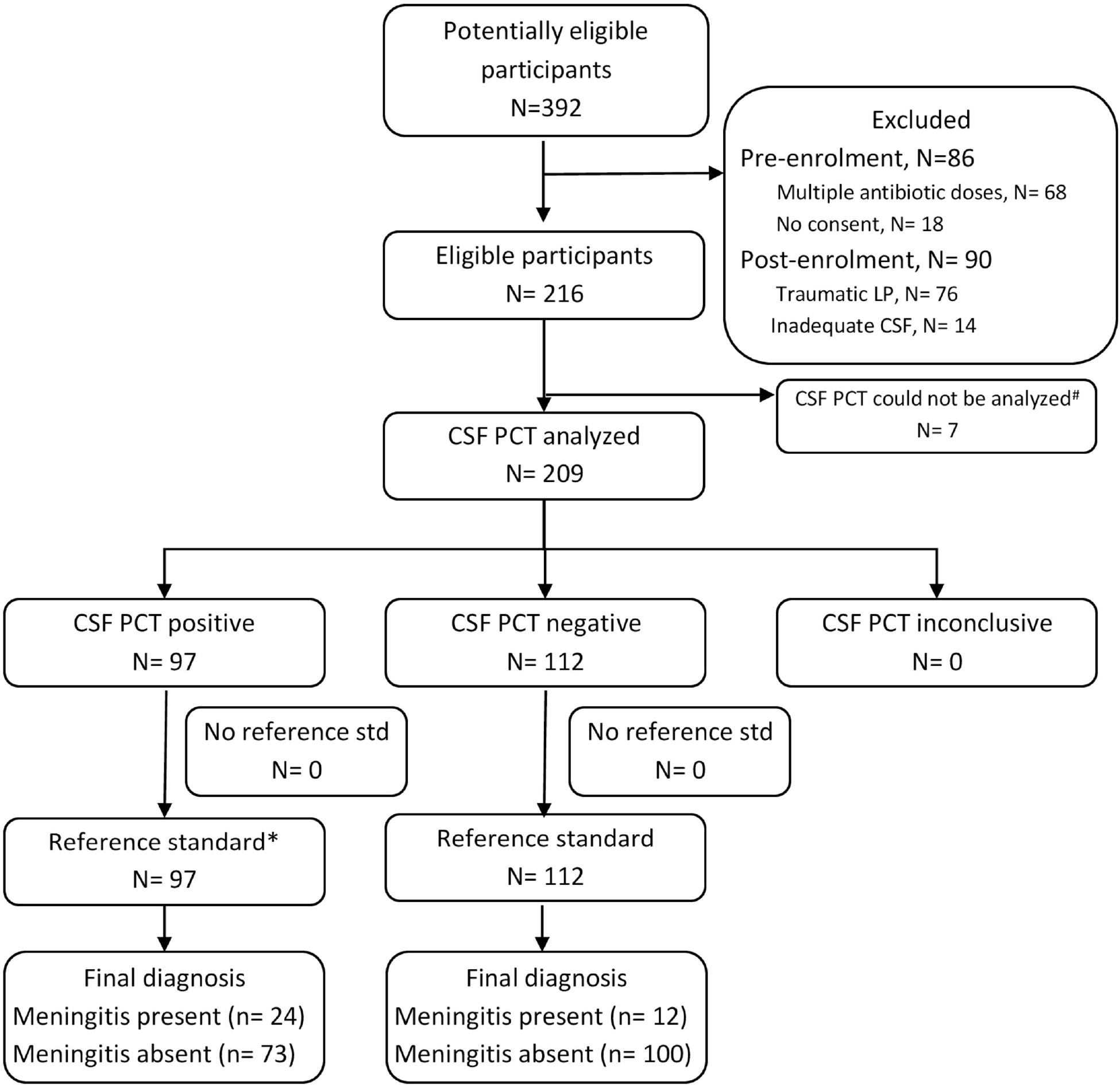
Flow diagram of study population. *In this flow diagram, reference standard is “definite or probable meningitis” ^#^ Of the 7 subjects whose CSF PCT could not be analysed, one was reference standard positive and 6 reference standard negative

The baseline characteristics were similar between the “definite” and “no definite meningitis” groups, barring encephalopathy, which was significantly commoner in the former (Table 1). We could not analyze CSF PCT in 7 samples due to logistic reasons, and plasma PCT in 69 neonates owing problems in drawing the extra blood sample.

**Table 1.** Comparison of characteristics between patients with definite meningitis and no definite meningitis

| Characteristics | Definite meningitis<br>(N = 18) | No definite<br>meningitis (N = 198) | P value |
| --- | --- | --- | --- |
| Birth weight, grams, median (1<sup>st</sup>, 3<sup>rd</sup> quartile) | 1297 (1051, 1692) | 1520 (1120, 2306) | 0.1 |
| Gestation, weeks, mean (SD) | 31.32 (2.83) | 32.7 (3.79) | 0.1 |
| Postnatal age, days, median (1<sup>st</sup>, 3<sup>rd</sup> quartile) | 6 (4, 10) | 6 (4, 15) | 0.8 |
| Apgar score at 5 minutes, median (1<sup>st</sup>, 3<sup>rd</sup> quartile) | 9 (8, 9) | 8 (7, 9) | 0.09 |
| Males, n (%) | 10 (55.5) | 128 (64.5) | 0.3 |
| Appropriateness for gestation status |  |  |  |
| AGA, n (%) | 16 (88.8) | 127 (64.1) | 0.078 |
| SGA, n (%) | 2 (11.1) | 64 (32.3) |  |
| LGA, n (%) | 0 | 7 (3.5) |  |
| PROM >18 h, n (%) | 2 (11.1) | 17 (8.6) | 0.8 |
| pPROM, n (%) | 4 (22.2) | 22 (11.1) | 0.2 |
| Clinical chorioamnionitis, n (%) | 1 (5.5) | 4 (2) | 0.4 |
| Foul smelling liquor, n (%) | 0 | 5 (2.5) | 0.5 |
| Seizures, n (%) | 3 (16.7) | 10 (5.0) | 0.06 |
| Encephalopathy, n (%) | 4 (22.2) | 10 (5.0) | **0.007** |
| Bulging anterior fontanelle, n (%) | 0 | 1 (0.5) | 0.8 |
| CSF PCT analysed, n (%) | 18 (100.0) | 191 (96.5) | 0.6 |
| Plasma PCT analysed, n (%) | 18 (100.0) | 129 (65.2) | 0.009 |

SD= standard deviation
AGA= appropriate for gestational age
SGA= small for gestational age
LGA= large for gestational age
PROM= prolonged rupture of membranes
pPROM= preterm premature rupture of membranes
CSF= cerebro-spinal fluid
PCT= Procalcitonin

The median (1^st^, 3^rd^ quartile) CSF PCT level was higher among neonates with definite compared to no definite meningitis [0.429 (0.123, 1.300) vs. 0.181 (0.119, 0.286) ng/ml respectively, p=0.028] (Table 2). Plasma PCT level and CSF: plasma PCT ratio did not differ significantly between the groups. The WBCC, neutrophil count, and protein concentration were significantly higher in the definite than the no definite meningitis group. The median (1^st^, 3^rd^ quartile) CSF PCT level was also higher among neonates with “definite or probable meningitis” compared to “no definite or probable meningitis” [0.245 (0.136, 0.675) vs. 0.170 (0.116, 0.28), p=0.01] (Table 3).

**Table 2.** Comparison of index tests between patients with definite meningitis and no definite meningitis

| Parameters | Definite meningitis<br>(N = 18) | No definite<br>meningitis (N = 198) | P value |
| --- | --- | --- | --- |
| CSF Procalcitonin, ng/ml | N=18<br>0.429 (0.123, 1.300) | N=191<br>0.181 (0.119, 0.286) | **0.028** |
| Plasma Procalcitonin, ng/ml | N=18<br>1.171 (0.275,<br>32.065) | N=129<br>0.459 (0.201, 1.905) | 0.09 |
| CSF: plasma Procalcitonin ratio | N=18<br>0.125 (0.049, 0.936) | N=129<br>0.319 (0.089, 0.748) | 0.3 |
| CSF total leukocyte count, cells per μL | 0 (0, 161) | 0 (0, 3.5) | **0.045** |
| CSF neutrophil count, cells per μL | 0 (0, 85) | 0 (0, 0) | **0.003** |
| CSF glucose, mg/dL | 65 (33, 82) | 56.5 (46, 70) | 0.8 |
| CSF protein, mg/dL | 126 (90, 187) | 92.5 (72.25, 124.5) | **0.003** |

All values are median (1<sup>st</sup>, 3<sup>rd</sup> quartile)
Wherever "N=" is not mentioned, it means samples of all patients in that cell were analyzed
CSF = cerebro-spinal fluid

**Table 3.** Comparison of Procalcitonin between patients with “definite or probable meningitis” and “no definite or probable meningitis”

| Parameters | Definite or probable meningitis (N = 37) | No definite or probable meningitis (N = 179) | P value |
| --- | --- | --- | --- |
| CSF Procalcitonin, ng/ml | N=36<br>0.245 (0.136, 0.675) | N=173<br>0.170 (0.116, 0.28) | **0.01** |
| Plasma Procalcitonin, ng/ml | N=32<br>0.561 (0.281, 5.292) | N=115<br>0.450 (0.196, 2.390) | 0.18 |
| CSF: plasma Procalcitonin ratio | N=32<br>0.249 (0.066, 0.808) | N=115<br>0.319 (0.087, 0.772) | 0.73 |

All values are median (1<sup>st</sup>, 3<sup>rd</sup> quartile)
CSF = cerebro-spinal fluid

The AUC of CSF PCT to diagnose definite meningitis was 0.656 (p=0.028) (Table 4). The optimal cut-off was 0.554 ng/ml. The AUC of CSF protein level was statistically significant (p=0.003), and that of neutrophil count neared statistical significance. Paired comparisons of the AUC of CSF PCT with other index tests did not show significant differences, though the difference between CSF PCT vs. CSF:plasma PCT ratio neared significance (p=0.053). The AUC of CSF PCT for diagnosing “definite or probable meningitis” was 0.635 (p=0.01). The optimal cut-off was 0.486 ng/ml. The difference between the AUC of CSF PCT and CSF:plasma PCT ratio was statistically significant (p-0.04). Based on the cut-off value of 0.2 ng/ml, CSF PCT had a sensitivity (95% CI) of 66.7% (50.4, 80.2), specificity of 57.8 (54.4, 60.6), LR^+^ of 1.58 (1.11, 2.04) and LR^-^ of 0.58 (0.33, 0.91).

**Table 4.** Area under the ROC curves of index tests for diagnosing meningitis

| **Outcome variable: definite meningitis** |  |  |  |  |  |
| --- | --- | --- | --- | --- | --- |
| **Index test** | **AUC** | **95% CI of AUC** | **P value of AUC** | **Cut-off based on Youden's index** | **P value of paired comparison of AUC of CSF Procalcitonin vs other index tests** |
| CSF Procalcitonin | 0.656 | 0.487, 0.826 | **0.028** | 0.554 | --- |
| Plasma Procalcitonin | 0.624 | 0.474, 0.775 | 0.09 | 4.87 | 0.61 |
| CSF: plasma Procalcitonin ratio | 0.430 | 0.273, 0.586 | 0.3 | 4.65 | 0.053 |
| CSF TLC | 0.612 | 0.456, 0.769 | 0.11 | 59 | 0.79 |
| CSF neutrophil count | 0.636 | 0.482, 0.791 | **0.05** | 22 | 0.93 |
| CSF glucose | 0.485 | 0.315, 0.656 | 0.8 | 19 | 0.22 |
| CSF protein | 0.709 | 0.588, 0.829 | **0.003** | 110 | 0.48 |
| **Outcome variable: definite or probable meningitis** |  |  |  |  |  |
| **Index test** | **AUC** | **95% CI of AUC** | **P value of AUC** | **Cut-off based on Youden's index** |  |
| CSF Procalcitonin | 0.635 | 0.525, 0.746 | **0.01** | 0.486 | -- |
| Plasma Procalcitonin | 0.578 | 0.465, 0.691 | 0.18 | 0.425 | 0.23 |
| CSF: plasma Procalcitonin ratio | 0.480 | 0.365, 0.595 | 0.7 | 1.91 | **0.04** |

ROC = receiver operator characteristics
AUC = area under the ROC curve
CSF = cerebro-spinal fluid

The median (1^st^, 3^rd^ quartile) CSF PCT level in subjects with LP performed at <72 hours (N=43) was higher than at ≥72 hours of life (N=166) [0.238 (0.149, 0.416) versus 0.170 (0.113, 0.286) ng/ml respectively, p=0.01]. The AUC of CSF PCT to diagnose “definite or probable meningitis” were similar at <72 hours versus ≥72 hours of life [0.68 (0.43, 0.94) versus 0.63 (0.51, 0.76) respectively]. CSF and plasma PCT values were correlated at <72 hours of life and at ≥72 hours of life [Spearman’s rho 0.54 (p=0.009) and 0.32 (p<0.001), respectively, (p value of difference between coefficients= 0.12)]. CSF and plasma PCT values were correlated among patients with “definite or probable meningitis” and those with no meningitis [rho= 0.47 (p= 0.007) and rho= 0.37 (p<0.001), respectively, (p value of difference between coefficients=0.29)].

## Discussion

The incidence of definite meningitis in our study population was 8.3%. To the best of our knowledge, this is the most extensive study on CSF PCT for NBM so far. CSF PCT, WBCC, neutrophil count, and protein were significantly higher in subjects with definite versus no definite meningitis, but plasma PCT and CSF: plasma PCT ratio were similar. Likewise, CSF PCT level was significantly higher in subjects with “definite or probable” meningitis, but plasma PCT level and CSF: plasma PCT ratio were not. CSF PCT did not have a high discriminatory value in our study, as the AUCs to diagnose definite meningitis and “definite or probable meningitis” were modest. On head-to-head comparisons between the AUC of CSF PCT and the other index tests, there were no significant differences. Using an *a priori* cut-off value, CSF PCT did not have good sensitivity, specificity, and LRs. Both CSF and plasma PCT values were higher at <72 hours of life than ≥72 hours. CSF PCT values were positively correlated with plasma PCT values.

Previous studies on neonates corroborate that CSF PCT values are significantly higher in patients with NBM.^22, 23^ However, the magnitude of effect in our study was substantially lower than the previous studies. Reshi et al. reported an AUC of 0.926 and Rajial et al. an AUC of 0.995 for CSF PCT. There could be several reasons for the differences. Reshi et al. recruited heavier babies (median 2.8 kg), and Rajial et al. more mature babies (> 34 weeks) than us. Neither research group included PCR as a reference standard criterion. Rajial et al. excluded neonates with any prior exposure to antibiotics, whereas we excluded only those exposed to multiple doses. Rajial et al. had a case-control diagnostic study design. Although Reshi et al. do not explicitly mention a two-gate, case-control design, but from the high proportion of meningitis (44.6%), it seems these groups were independently recruited. The cut-off values in these studies, 0.33 ng/ml and 0.2 ng/ml respectively, were derived and validated in the same data set. In diagnostic test studies, a case-control design and validation of cut-off values on the derivation data set can inflate the estimates of diagnostic accuracy.^25^

There are a few studies on CSF PCT in post-neonatal children. Among children with suspected meningitis, Shokrollahi et al. found significantly higher CSF PCT levels in the bacterial meningitis group compared to aseptic meningitis [1.55 ± 1.19 versus 0.39 ± 0.33 ng/ml, p <0.001].^26^ Among symptomatic children aged 28 days to 14 years, Sanaei Dashti et al. reported that CSF PCT had a sensitivity of 75% and specificity of 47.4% to diagnose definite or probable meningitis.^27^ In another study on children (mean ∼5 y), the median (95% CI) CSF PCT level was higher in bacterial meningitis [1 (0.2, 2.56)] than viral meningitis [0.05 (0.05, 0.05), p <0.05] and non-meningitis [0.07 (0.05, 0.1), p <0.05].^28^ Like our study, the AUC of CSF PCT was higher than serum PCT (0.76 vs. 0.68, p<0.05). On the contrary, Sanaei Dashti et al. did not find significant differences between CSF and serum PCT.^27^

There are several studies on serum PCT for the diagnosis of bacterial meningitis in children. Among children aged 3 m-15 y, serum PCT (cut-off 0.5 ng/ml) had a sensitivity and specificity of 95.45% and 84.61% with an AUC of 0.99.^29^ Among children (4 months-14 years) mean (SD) serum PCT (ng/ml) was higher in bacterial meningitis versus aseptic meningitis [p<0.001].^30^ Vikse et al. combined 8 studies on serum PCT to diagnose bacterial meningitis in post-neonatal children.^18^ Threshold values varied from 0.2 ng/ml to 3.3 ng/ml. The pooled sensitivity (95% CI) was 0.96 (0.92, 0.98) and pooled specificity 0.89 (0.86, 0.92).

The source of PCT in the CSF in meningitis is unclear, but it could be from the plasma or inflamed brain tissue or activated leukocytes in the CSF. Banks states that inflammatory cytokines in the blood may disrupt the blood-brain barrier, cytokines may cross from the blood to the CSF and vice versa, and the blood-brain barrier may itself produce cytokines.^31^ Meningitis is associated with a higher degree of bacteremia.^32^ A more intense inflammatory response in response to a higher degree of bacteremia may explain both the elevated levels of plasma and CSF PCT observed in bacterial meningitis. Though we found higher plasma PCT levels at <72 hours (as expected), the similarity in coefficients of correlation between CSF and plasma PCT at <72 hours versus ≥72 hours suggests that the movement of PCT from the blood to CSF contributes to CSF PCT.

In health, minimal PCT is produced by cells in the thyroid, lung, and intestines but no other tissues. However, in an animal model of sepsis, calcitonin messenger RNA was expressed by brain tissue, which may be a source of PCT in meningitis.^33^ It is not clear whether activated leucocytes in the CSF can also produce PCT. In *in vitro* experiments, placental macrophages upregulate PCT expression following lipopolysaccharide stimulation.^34^ No calcitonin gene-related peptide expression (CGRP) is observed in leukocytes of septic patients with high levels of PCT, but cytokine-induced expression of PCT and CGRP from adipocytes is observed.^35^ In contrast, Balog et al. found that TNF-alpha could stimulate increased PCT production from leukocytes.^36^

The strengths of our study were a large sample size, a single-gate design, blinding, and definition of a cut-off value *a priori*. The limitations included a small number of reference standard positive subjects, lack of information regarding the diagnostic spectrum of non-meningitis patients, inability to prove viral meningitis, and missing the plasma PCT test on several subjects.

There is a need for further methodologically sound, adequately powered studies on PCT and other inflammatory biomarkers in the CSF to diagnose NBM. Further research is required into the source of PCT in the CSF and why serum PCT increases in meningitis over and above the increase in sepsis.

## Conclusions

In our study, there is an association between bacterial meningitis and CSF PCT values. However, we did not find sufficient separation of values of CSF PCT between meningitis and non-meningitis groups. Hence, CSF PCT could not reliably discriminate between meningitis and no meningitis.

## Data Availability

Data will be made available on written request

## Contributorship statement

Sourabh Dutta made substantial contributions to the conception and design of the work, acquisition, analysis and interpretation of data AND drafted the work and revised it critically for important intellectual content AND approved the final version to be published AND agrees to be accountable for all aspects of the work

Naresh Sachdeva made substantial contributions to the design of the work, acquisition, analysis, and interpretation of data AND revised the work critically for important intellectual content AND approved the final version to be published AND agrees to be accountable for all aspects of the work.

Vilay Sanger made substantial contributions to the design of the work, and acquisition of data AND revised the work critically for important intellectual content AND approved the final version to be published AND agrees to be accountable for all aspects of the work.

Arnab Pal made substantial contributions to the design of the work, acquisition, analysis, and interpretation of data AND revised the work critically for important intellectual content AND approved the final version to be published AND agrees to be accountable for all aspects of the work.

Pallab Ray made substantial contributions to the concept and design of the work, acquisition, analysis, and interpretation of data AND revised the work critically for important intellectual content AND approved the final version to be published AND agrees to be accountable for all aspects of the work.

## What is already known on this topic

- Elevated *serum/plasma* Procalcitonin is associated with bacterial meningitis among adults, children, and young infants.
- Elevated *cerebrospinal fluid* (CSF) Procalcitonin is associated with bacterial meningitis among adults and children in the post-neonatal age-group.
- There is a paucity of studies regarding CSF and serum/plasma Procalcitonin for the diagnosis of neonatal bacterial meningitis.

## What this study adds

- Cerebro-spinal fluid Procalcitonin and plasma Procalcitonin do not have sufficient ability to discriminate between meningitis and no meningitis among neonates with clinical sepsis.
- The diagnostic performance of cerebro-spinal fluid Procalcitonin is not superior to CSF leukocyte count and biochemistry.
- There is a high correlation between CSF and plasma Procalcitonin values; the coefficients of correlation are similar between less than 72 hours versus ≥ 72 hours of life and between neonates with and without meningitis.

## Acknowledgments

The authors acknowledge the essential contributions of Prof Anuradha Chakraborty, Head of the Department of Experimental Medicine and Biotechnology at PGIMER, Chandigarh, India, who helped with the study’s design and establishing the molecular methods.

